# A pilot investigation of enteric pathogens and stunting and malnutrition using a combination of hospital surveillance and a birth cohort of children in Dili, Timor-Leste

**DOI:** 10.1101/2023.02.21.23286223

**Authors:** Danielle M. Cribb, Nevio Sarmento, Almerio Moniz, Nicholas S. S. Fancourt, Kathryn Glass, Anthony D. K. Draper, Joshua R. Francis, Milena M. Lay dos Santos, Endang Soares da Silva, Benjamin G. Polkinghorne, Virginia de Lourdes da Conceição, Feliciano da Conceição, Paulino da Silva, Joanita Jong, Martyn D. Kirk, Samantha Colquhoun

## Abstract

**Background:** Enteric pathogens contribute to child malnutrition in low-to-middle-income countries. In Timor-Leste, there has been limited study of this relationship.

**Methodology/Principal findings:** We investigated enteric disease, stunting, and malnutrition in children in Dili, Timor-Leste (July 2019 – October 2020). Sixty infants received up to four home visits. We collected faecal samples, demographics, anthropometrics, food and water sources, feeding practices, and animal husbandry details. For 160 children admitted to Hospital Nacional Guido Valadares with clinical diagnosis of severe diarrhoea or severe acute malnutrition (SAM), we collected faecal samples, diagnostics, and anthropometrics. Faeces were tested by polymerase chain reaction (PCR) using the BioFire^®^ FilmArray^®^ Gastrointestinal Panel. Descriptive analyses and regression analyses were conducted using R.

We detected enteric pathogens in 68.8% (95% confidence interval [CI] 60.4–76.2%) of infants, 88.6% of children with SAM (95% CI 81.7–93.3%), and 93.8% of children with severe diarrhoea (95% CI 67.7– 99.7%). Diarrhoeagenic *Escherichia coli* and *Campylobacter* spp. were most frequently detected. A median weight-for-height Z-score (WHZ) of -1.1 (interquartile range [IQR] -2.0 to -0.1) for infants across all home visits, -3.9 (IQR -4.7 to -3.0) for children with SAM and -2.6 (IQR -4.2 to -1.1) for children with severe diarrhoea was observed.

The most common admission diagnosis was SAM (88.1%, 95% CI 81.7–92.5%). Hospitalised children had increased odds of *Shigella*/enteroinvasive *E. coli* (OR 11.4, 95% CI 2.8–47.3) and *Giardia lamblia* detection (OR 31.7, 95% CI 3.6–280) compared with infants. For home visits, bottle feeding was associated with increased odds of pathogen detection (OR 8.2, 95% CI 1.1–59.7).

**Conclusions/Significance:** We detected high prevalence of enteric pathogens and signs of malnutrition in children in Dili. Our pilot is proof of concept for a study to fully explore the risk factors and associations between enteric pathogens and malnutrition in Timor-Leste.

**Author summary:** Enteric pathogens are a contributing factor to child malnutrition in low-to-middle-income countries. This study investigated the relationship between enteric disease, malnutrition, and stunting in children in Dili, Timor-Leste. Sixty infants received up to four home visits and 160 children were admitted to the hospital with a clinical diagnosis of severe diarrhoea or severe acute malnutrition. Faecal samples were collected and tested for enteric pathogens and risk factors were explored. The study found that enteric pathogens were detected in most infants and children with malnutrition and diarrhea. The most commonly detected pathogens were diarrheagenic *Escherichia coli* and *Campylobacter* spp. Most children exhibited signs of malnutrition from infancy, and hospitalised children had a higher likelihood of having certain enteric pathogens. Feeding practices, water use, and animal cohabitation may pose risks for enteric pathogen infection. The results suggest that enteric pathogens are a significant problem in Dili, Timor-Leste, and further research is needed to explore the complexity of risk factors.

## Introduction

Childhood undernutrition is a major challenge for many countries and is associated with high levels of morbidity and mortality (1). In 2020, 22.0% of children aged under five years worldwide suffered from stunting (low height for age) and 6.7% suffered from wasting (low weight for height) (2). Children in low- and middle-income countries (LMICs) are disproportionately impacted by stunting (38.8%) and wasting (12.9%) (3). Undernutrition can lead to cognitive, physical, and metabolic developmental issues, potentially reducing physical and intellectual ability and economic productivity in adulthood (1).

Socioeconomic and environmental factors contribute to child undernutrition, including poor maternal health and education, early termination of breastfeeding, low household wealth, and unimproved sanitation (3, 4). There is growing evidence for an association between infection with enteric bacteria, viruses, and parasites and child stunting and wasting, particularly in LMICs (5, 6). Enteric bacterial pathogens contracted early in life may damage the lining of the small and large intestines, reducing the absorption of nutrients from food and promoting chronic inflammation, leading to undernutrition and a compromised immune system (5, 6). Such enteric pathogens potentially involved in stunting and malnutrition include enterotoxigenic *Escherichia coli* (ETEC), *Shigella*, and *Campylobacter* species (7-9).

*Campylobacter* is the most common cause of foodborne bacterial gastroenteritis globally, causing an estimated 96 million cases and >21,000 deaths annually (10). In high-income settings, infections are often sporadic in nature and largely attributed to consuming undercooked chicken (11). In low-income settings, *Campylobacter* is frequently endemic and while still largely attributed to poultry, infection determinants are from more varied environmental and zoonotic sources (11, 12). Recent studies have further implicated *Campylobacter* in child stunting and malnutrition. A Peruvian cohort exhibited poor weight gain with infection and marginal poor linear growth with symptomatic infection (7, 9). Similarly, 84.9% of the multisite Malnutrition and Enteric Disease Study (MAL-ED) cohort presented with a *Campylobacter-*positive faecal sample by one year of age, with those diagnosed with multiple *Campylobacter* infections being more likely to exhibit poor linear growth (12).

Timor-Leste is one of the lowest-income countries in Asia and has a population of 1.34 million circa 2022 (13, 14). The 2016 Demographic and Health Survey (DHS) estimated that the proportion of children with stunting was 46.5%, wasting 24.2% and underweight (low weight for age) 40.4% (15). Diarrhoeal disease is prevalent in Timor-Leste with 10.7% of children represented in the DHS presenting with diarrhoea within the two weeks preceding the survey, peaking at 17.8% in children aged 12-23 months (16).

We conducted a pilot investigation to examine the feasibility of a birth cohort and hospital-based surveillance study in Timor-Leste. We aimed to i) estimate the proportion of children from either a birth cohort or hospital admissions with severe acute malnutrition (SAM) and/or severe diarrhoea who tested positive for enteric pathogens on stool, ii) estimate the proportion of children from a birth cohort who were malnourished or stunted at any time during the first 12 months of life, iii) quantify malnourishment in children admitted to hospital with SAM and/or severe diarrhoea, iv) describe the social, environmental, food safety, and food security issues associated with enteric pathogen infections in Dili, Timor-Leste, and v) support capacity building in research and laboratory skills. This investigation formed the basis of a larger study to understand the impact of enteric pathogens on stunting and malnutrition.

## Methods

This study was carried out at the national referral hospital in Dili, Timor-Leste, Hospital Nacional Guido Valadares (HNGV). HNGV has a catchment population of 500,000 people with approximately 4,500 births per year.

### Birth cohort

#### Study setting and recruitment

We recruited newborns into the longitudinal birth cohort between July and September 2019. Newborn infants were eligible for inclusion if they were born at ≥38 weeks gestation at HNGV and if they were reported to live within a 10-kilometre radius of HNGV. The local research team approached parents in the maternity unit at HNGV after delivery and prior to discharge. Parents provided informed consent to enrol their infants. Home visits occurred when infants were approximately one, four, seven, and 12 months of age.

#### Equipment and questionnaire

The local study team were trained to use a calibrated portable baby scale and a standardised mat for measuring weight (Seca 334 Baby Scales) and height (Seca 210 Mobile Measuring Mat for Babies and Toddlers). All infants were weighed and measured using a standardised procedure. In addition, the team interviewed parents using a standardised questionnaire at each visit to investigate feeding practices, household size, water, sanitation, and hygiene (WASH) practices and accessibility, and household animal ownership and containment (16). All data were collected on an android tablet using Research Electronic Data Capture (REDCap) (17, 18), synchronised online, and stored on a secure server hosted at the Australian National University (ANU).

#### Specimen collection and testing

Faecal samples were collected in sample pots from all infants at the time of the field visit or in the following days and transported to the lab in an insulated United Nations Children’s Fund container. Parents and/or guardians were provided with instructions in their native language, Tetum to ensure samples were correctly collected and stored prior to transport. Samples were transported to the National Health Laboratory (NHL) for polymerase chain reaction (PCR) testing and culture for *Campylobacter, Shigella or Salmonella* isolates. Animal faecal samples were collected from the environment proximal to homes where a positive *Campylobacter, Shigella* or *Salmonella* infant faecal sample was detected. Samples were transported to the Timor-Leste Diagnostics Laboratory for Animal Health (DLAH) for preparation and isolate culture on appropriate selective media.

### Hospital-based surveillance

#### Study setting and recruitment

We recruited children admitted to HNGV with SAM, diarrhoea with severe dehydration (DWSD), or dysentery between October 2019 to July 2020 to investigate enteric pathogens in children with these conditions in Dili. Children were eligible for inclusion if they were aged less than five years at time of hospital admission, if they resided within Dili, and if they met the adapted World Health Organization (WHO) criteria for diagnosis of SAM, DWSD and/or severe dysentery (19).

#### Specimen and information collection

Participants were identified for eligibility by paediatricians during the daily paediatric ward round. Following informed consent from parents or guardians, ward nurses collected faecal samples from eligible children and the study team recorded vital statistics and demographic characteristics from the medical notes and transported the samples to the NHL for PCR testing.

### Pathogen detection

The NHL used a BioFire^®^ FilmArray^®^ Gastrointestinal Panel PCR assay to test all faecal samples from birth cohort and hospitalised children to detect 22 enteric pathogens and parasites (20). Samples positive by PCR for selected enteric bacteria (*Campylobacter, Salmonella* and *Shigella*/EIEC) were reflex cultured on appropriate selective media.

### Data analysis

All data were cleaned and analysed using R statistical software (21). We converted anthropometric measures to weight-for-height (WHZ), height-for-age (HAZ), and weight-for-age (WAZ) Z scores and mapped these against the WHO child growth standards (22, 23) using R package “zscorer” (24). We performed Fisher’s exact and Chi-square tests to analyse differences between groups and visits. We calculated odds ratios (OR) using generalised estimating equations (GEE) to account for repeated observations with the R package “geepack”, where appropriate (25). GEE were used to analyse risk factors in the birth cohort against the outcomes of i) pathogen detection and ii) moderate wasting, defined as a WHZ of -2 or below (26). Additionally, GEE were used to analyse associations between hospitalised SAM cases and non-hospitalised children for pathogen detections, demographics and faecal consistency. GEE models used an exchangeable correlation structure and binomial distribution with logistic link. We used the “arsenal” package to calculate generalised linear models, where appropriate (27). A *p* value of *p* < 0.05 was considered significant for Fisher’s and Chi-square tests, and an OR with confidence intervals (CI) that did not cross over one was considered significant for GEE.

### Ethics

The Timor-Leste National Institute of Health-Research Ethics & Technical Committee (reference 462/MS-INS/GDE/IV/2019) for the Timor-Leste Instituto Nacional de Saúde (INS), and the Australian National University Ethics Committee (reference 2019/221) provided ethical approval. The field study team were trained in Good Clinical Practice for research by the project lead (28). All parents or legal guardians provided informed consent prior to study enrolment.

## Results

### Birth cohort

We recruited 32 male and 28 female infants into the birth cohort. Only 53.3% (32/60) were retained until the final home visit. At birth, the median WHZ was 0.9 (interquartile range [IQR] -0.5 – 1.6), with no scores calculated for four newborns with lengths below 45cm (S1 Table). The median WHZ over all four home visits decreased to -1.1 (IQR -2.0 to -0.1). Of the 32 infants who were weighed and measured at all four visits, 31.3% (10/32) exhibited poor weight gain (WHZ of ≤ -3.0) on at least one visit. While the birth cohort tracked the WHO 50^th^ percentile for height-for-age, weight-for-age and weight-for-height tracked between the 15^th^ and 25^th^ percentiles (**Error! Reference source not found**.).

We collected 141 infant faecal specimens for PCR testing from 161 home visits (87.6% sample collection rate), with at least one faecal sample collected from 45/60 (75.0%) of participants. We detected at least one enteric pathogen in 68.8% (97/141; 95%CI 60.4%–76.2%) of samples, and of these 63.9% (62/97; 95% CI 53.5%–73.2%) were positive for multiple pathogens.

We observed significant differences in pathogen occurrence between home visits, with the number of pathogens detected within the cohort and the number of multiple pathogen detections in individual infants generally increasing with age (*p* < 0.05; **Table 1**). Twenty-two study participants had samples PCR tested at each visit, with each having at least one positive result for an enteric pathogen and 40.9% (9/22) positive at every visit. There was a weak relationship between WHZ and the number of pathogens detected per sample at each visit (**Fig 2**).

**Table 1:** Number and proportion of pathogens detected by multiplex PCR at each birth cohort home visit and stool presentation of samples, Dili, Timor-Leste, 2019–2020.

| Pathogen | Visit 1<br>n (%) | Visit 2<br>n (%) | Visit 3<br>n (%) | Visit 4<br>n (%) | Total<br>n (%) |
| --- | --- | --- | --- | --- | --- |
| **Bacteria** |  |  |  |  |  |
| *Campylobacter* spp.* | 3 (6.7) | 1 (2.9) | 3 (8.3) | 8 (30.8) | **15 (10.6)** |
| *Clostridioides difficile* | 1 (2.2) | 4 (11.8) | 4 (11.1) | 4 (15.4) | **13 (9.2)** |
| *Salmonella* | 0 (0) | 0 (0) | 4 (11.1) | 0 (0) | **4 (2.8)** |
| Shiga toxin-producing *Escherichia coli* (STEC) | 0 (0) | 0 (0) | 0 (0) | 2 (7.7) | **2 (1.4)** |
| Enteroaggregative *E. coli* | 11 (24.4) | 21 (61.8) | 30 (83.3) | 15 (57.7) | **77 (54.6)** |
| Enteropathogenic *E. coli* | 3 (6.7) | 10 (29.4) | 21 (58.3) | 15 (57.7) | **49 (34.8)** |
| Enterotoxigenic *E. coli* lt/st | 1 (2.2) | 4 (11.8) | 12 (33.3) | 5 (19.2) | **22 (15.6)** |
| *Shigella*/Enteroinvasive *E. coli* | 1 (2.2) | 0 (0) | 1 (2.8) | 0 (0) | **2 (1.4)** |
| *Plesiomonas shigelloides* | 0 (0) | 0 (0) | 1 (2.8) | 0 (0) | **1 (0.7)** |
| *Vibrio* spp. † | 1 (2.2) | 0 (0) | 4 (11.1) | 0 (0) | **5 (3.5)** |
| **Viruses** |  |  |  |  |  |
| Adenovirus F40/41 | 0 (0) | 0 (0) | 2 (5.6) | 4 (15.4) | **6 (4.3)** |
| Norovirus GI/GII | 0 (0) | 3 (8.8) | 7 (19.4) | 2 (7.7) | **12 (8.5)** |
| Rotavirus A | 0 (0) | 3 (8.8) | 2 (5.6) | 4 (15.4) | **9 (6.4)** |
| Sapovirus | 0 (0) | 0 (0) | 4 (11.1) | 0 (0) | **4 (2.8)** |
| **Parasites** |  |  |  |  |  |
| *Cryptosporidium* spp. | 0 (0) | 0 (0) | 0 (0) | 3 (11.5) | **3 (2.1)** |
| *Giardia lamblia* | 0 (0) | 0 (0) | 1 (2.8) | 0 (0) | **1 (0.7)** |
| **Total samples** | **45** | **34** | **36** | **26** | **141** |
| * *Campylobacter* spp. includes *C. jejuni*, *C. coli*, and *C. upsaliensis*. † *Vibrio* spp. includes *V. parahaemolyticus*, *V. vulnificus*, and *V. cholerae*.<br>N.B. percentages and counts do not equal total samples due to multiple pathogen detections in some samples. |  |  |  |  |  |

**Fig 1.**
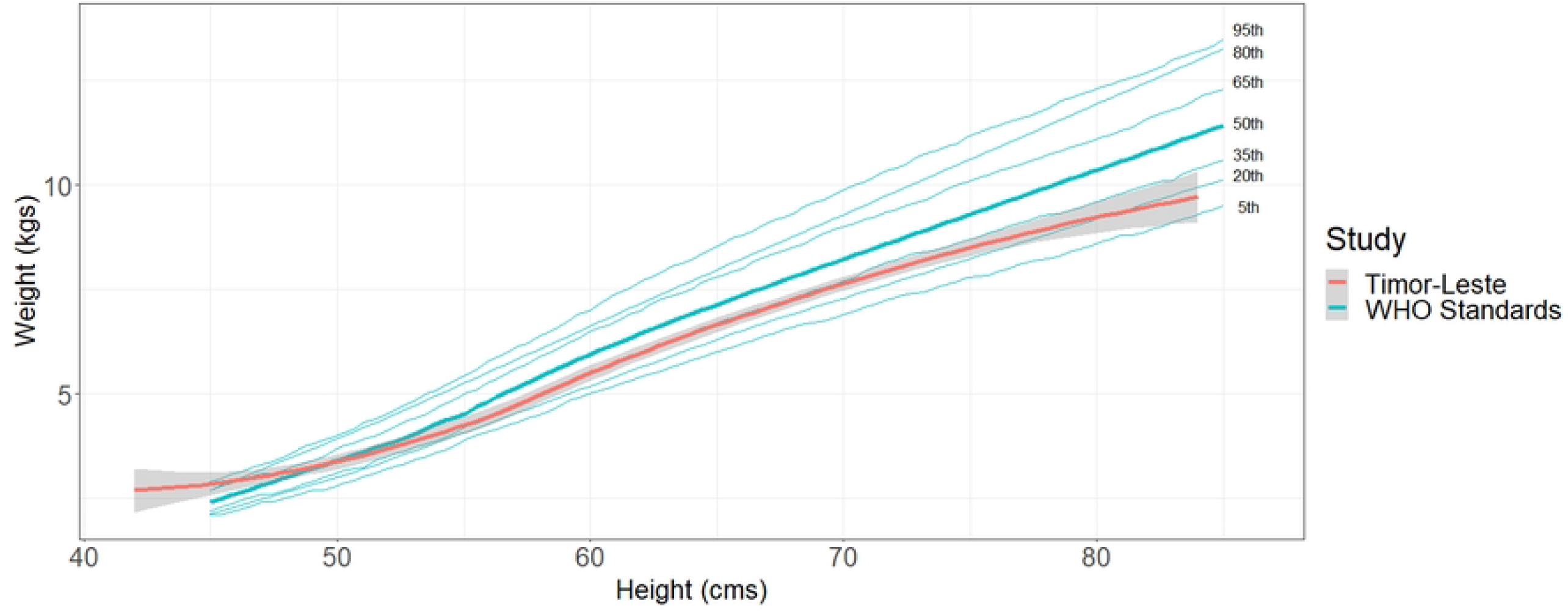
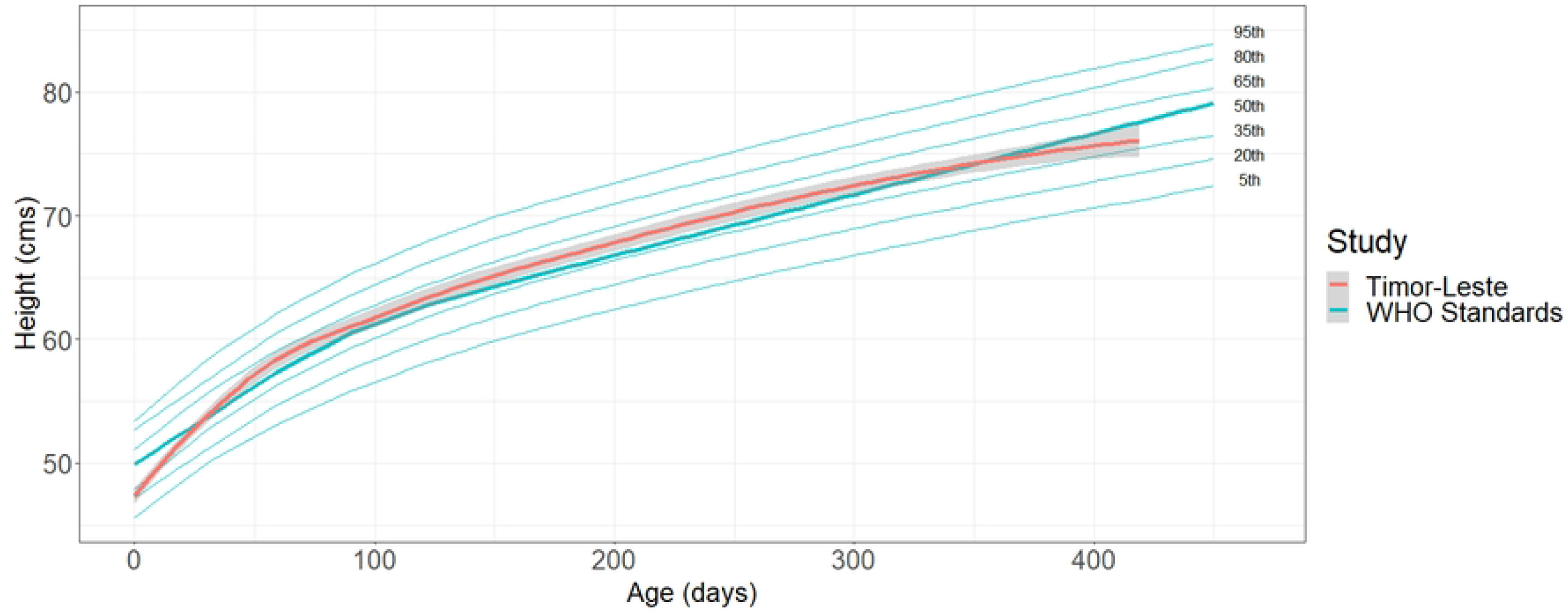
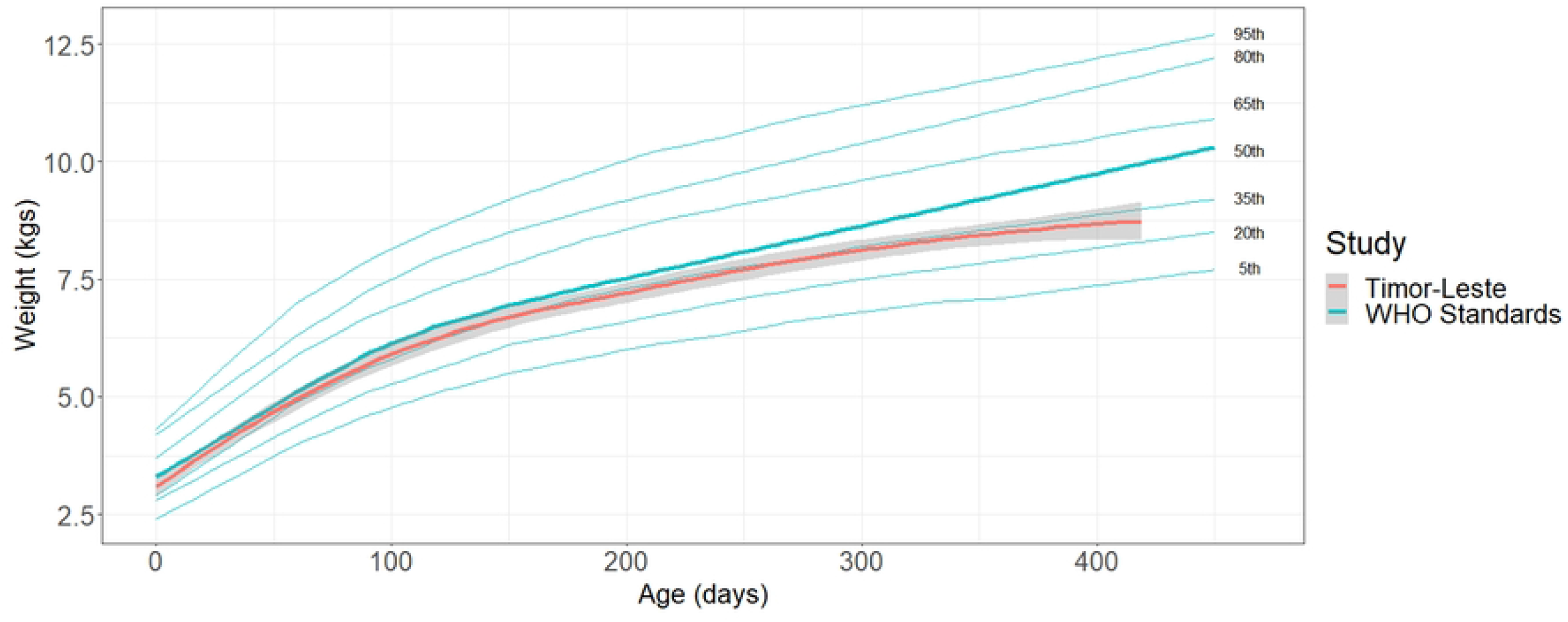
(a) Height-for-age World Health Organization standards compared with Timor-Leste birth cohort, 2019–2020; (b) Weight-for-age WHO standards compared with Timor-Leste birth cohort, 2019–2020; (c) Weight-for-height WHO standards compared with Timor-Leste birth cohort, 2019–2020. WHO percentiles are annotated on figure. 50^th^ percentile for WHO standards is presented in bold. Grey shading represents the 95% confidence interval for the Timor-Leste birth cohort.

**Fig 2.**
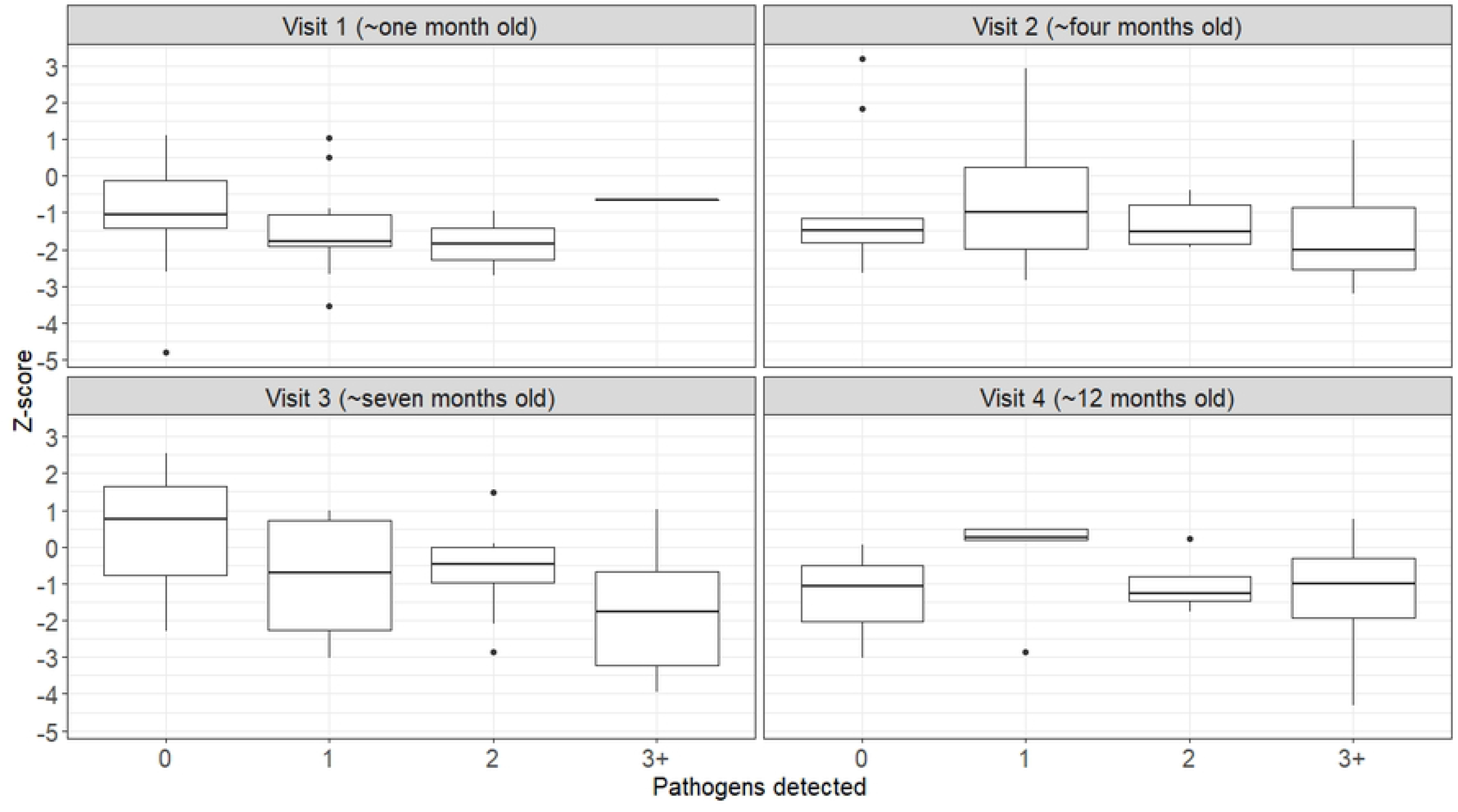
Weight-for-height Z-score by number of pathogens detected for each birth cohort visit, Dili, Timor-Leste, 2019–2020.

Diarrhoeagenic *E. coli* (DEC) strains (Shiga toxin-producing *E. coli* [STEC], EPEC, EAEC, and ETEC) were the most common pathogens detected. Six infants were PCR positive for DEC at all four visits, with EAEC detected in 96.0% (23/24) of their faecal samples, indicating possible chronic carriage. Fifteen samples were PCR positive for *Campylobacter* spp., four samples of *Salmonella*, and two samples of *Shigella*/enteroinvasive *E. coli* (EIEC) across the entire cohort. During the first home visits, three samples collected from animal faeces were successfully cultured and *Campylobacter* spp. isolated by the DLAH team. No isolates were cultured from human clinical samples.

Full descriptive results and ORs for these analyses are available in S2 Table and S3 Table. Compared to children aged <3 months, we found that those aged 3–<6 months (OR 5.4, 95% CI 2.1–14.3), 6–<9 months (OR 18.1, 95% CI 4.8–68.8), and ≥12-months (OR 12.1, 95% CI 3.1–47.0) had higher odds of pathogen detection, i.e., the odds of pathogen detection increased with age. We controlled for age in analysis of all additional risk factors. We found that exclusive bottle feeding was associated with increased odds of enteric pathogen detection (OR 8.2, 95% CI 1.1–59.7) compared with breastfeeding. No other factors were found to be statistically significant.

For the outcome of wasting, controlled for age, we found that households with five or fewer members were associated with lower odds of wasting (OR 0.2, 95% CI 0.1–1.0) compared to those with 6–10 household members. No other factors were significant.

### Hospital-based surveillance

We recruited 160 children in hospital-based surveillance. One child was excluded as they were aged above five years. We included 159 children in descriptive analyses including those that were missing anthropometric data including gender (n=2), middle-upper arm circumference (n=5), and height (n=1). Females represented 52.9% (83/157) of participants with known gender. We received PCR results from 148 participants as two patients were discharged before specimen collection and nine were missing laboratory PCR data.

The mean age at admission was 17.7 months (range 1.0–55.7 months, standard deviation (*SD*) ±11.4). We diagnosed 140 (88.1%, 95% CI 81.7–92.5%) children with SAM and 19 (11.9%, 95% CI 7.5–18.3%) with severe diarrhoeal disease (13 with DWSD, and six with dysentery). The median WHZ for those diagnosed with SAM (−3.9, IQR -4.7 to -3.0) was lower than those with severe diarrhoea (−2.6, IQR -4.2 to -1.1). Six WHZ were not calculated for SAM participants due to unknown gender (n=2), unknown height (n=1) or a height below 45cm (n=3).

Of the 148 participants with complete multiplex PCR results, 88.6% (117/132; 95% CI 81.7–93.3%) of SAM participants were positive for at least one enteric pathogen, with 73.5% (97/132; 95% CI 65.0– 80.6%) positive for multiple pathogens. Of those diagnosed with severe diarrhoea, 93.8% (15/16; 95% CI 67.7–99.7%) were positive for at least one enteric pathogen and all but one were positive for multiple pathogens (14/16; 87.5%, 95% CI 60.4–97.8%). We detected EAEC and/or EPEC in 81.8% (9/11) of loose faecal samples. The age at admission for participants varied between the type of pathogen detected, with viral pathogens detected in admissions with a mean age of 13.8 months (*SD* ±8.1), bacteria at 17.8 months (*SD* ±11.2), and parasites at 19.3 months (*SD* ±11.7). There were no statistically significant differences for age at admission by pathogen group between SAM and severe diarrhoea cases (*p* > 0.05) (S4 Table).

The mean number of pathogens detected per individual was 2.8 (range 0–7). The most detected pathogens were DEC species followed by *Campylobacter* spp., *Shigella/*EIEC, and *Giardia lamblia* (**Table 2**). Two *Campylobacter* spp. and two *Salmonella* isolates were cultured.

**Table 2:** Hospital-based surveillance data of diagnosis-specific pathogen detections and total pathogen detections from PCR testing, Dili, Timor-Leste, 2019-2020.

| Pathogen | Pathogen detections in SAM cases<br>*n* (%) | Pathogen detections in severe diarrhoea<sup>‡</sup><br>cases<br>*n* (%) | All pathogen detections<br>*n* (%) |
| --- | --- | --- | --- |
| **Bacteria** |  |  |  |
| *Campylobacter* spp.* | 33 (25.0) | 3 (18.8) | 36 (24.3) |
| *Clostridioides difficile* | 6 (4.5) | 0 (0.0) | 6 (4.1) |
| *Plesiomonas shigelloides* | 1 (0.8) | 0 (0.0) | 1 (0.7) |
| *Salmonella* | 4 (3.0) | 1 (6.3) | 5 (3.4) |
| Shiga toxin-producing *Escherichia coli* (STEC) | 4 (3.0) | 0 (0.0) | 4 (2.7) |
| Enteroaggregative *E. coli* | 84 (63.6) | 10 (62.5) | 94 (63.5) |
| Enteropathogenic *E. coli* | 70 (53.0) | 10 (62.5) | 80 (54.1) |
| Enterotoxigenic *E. coli* lt/st | 41 (31.1) | 4 (25.0) | 45 (30.4) |
| *E. coli* O157 | 2 (1.5) | 0 (0.0) | 2 (1.4) |
| *Shigella*/Enteroinvasive *E. coli* | 23 (17.4) | 7 (43.8) | 30 (20.3) |
| *Vibrio* spp.† | 5 (3.8) | 3 (18.8) | 8 (5.4) |
| **Viruses** |  |  |  |
| Adenovirus F40/41 | 3 (2.3) | 1 (6.3) | 4 (2.7) |
| Astrovirus | 6 (4.5) | 1 (6.3) | 7 (4.7) |
| Norovirus GI/GII | 12 (9.1) | 1 (6.3) | 13 (8.8) |
| Rotavirus A | 6 (4.5) | 1 (6.3) | 7 (4.7) |
| Sapovirus | 13 (9.8) | 1 (6.3) | 14 (9.5) |
| **Parasites** |  |  |  |
| *Cryptosporidium* spp. | 17 (12.9) | 1 (6.3) | 18 (12.2) |
| *Giardia lamblia* | 31 (23.5) | 2 (12.5) | 33 (22.3) |
| *Cyclospora cayetanensis* | 12 (9.1) | 0 (0.0) | 12 (8.1) |
| *Entamoeba histolytica* | 0 (0.0) | 1 (6.3) | 1 (0.7) |
| **Total samples** | **132** | **16** | **148** |
| * *Campylobacter* spp. includes *C. jejuni*, *C. coli*, and *C. upsaliensis*. † *Vibrio* spp. includes *V. parahaemolyticus*, *V. vulnificus*, and *V. cholerae*. ‡Severe diarrhoea includes diarrhoea with severe dehydration and dysentery diagnoses. PCR = polymerase chain reaction test. SAM = severe acute malnutrition. N.B. percentages and counts do not equal total samples due to multiple pathogen detections in some samples. |  |  |  |

### Comparing hospitalised and non-hospitalised children

Hospitalised children had increased odds of both *Shigella*/EIEC (OR 11.4, 95% CI 2.8–47.3) and *Giardia lamblia* infection (OR 31.7, 95% CI 3.6–280) compared with the non-hospitalised birth cohort (S5 Table).

## Discussion

Infants and young children from a birth cohort and hospital-based surveillance study exhibited high enteric pathogen loads and signs of wasting and undernutrition from the first months of life. Ninety percent of children admitted to hospital for treatment of DWSD, dysentery, or SAM were positive for at least one enteric pathogen and 70% of infants visited at home also returned a positive PCR test for at least one enteric pathogen over the 12-month study. A high proportion of samples across both study arms had multiple pathogens recovered. Pathogen detection generally increased with age within the birth cohort, likely influenced by exposure to new nutrition sources and environmental factors. While the mechanisms of child malnutrition remain complex, our pilot study has demonstrated the ability to conduct longitudinal research in Timor-Leste to explore and understand the relationship between enteric pathogens and malnutrition.

We detected a variety of enteric pathogens in children regardless of the presence of wasting and diarrhoeal disease. Across symptomatic and asymptomatic faecal samples, *Campylobacter* spp., DEC strains EAEC, EPEC, and ETEC, *Shigella*/EIEC, norovirus, *Giardia*, and *Cryptosporidium* were most prevalent. This is consistent with results from the MAL-ED study (29) and the Global Enteric Multicenter Study (GEMS)(30). Diarrhoeal faecal samples in our study frequently had DEC strains present, suggesting these may be driving disease presentation in combination with non-pathogenic factors. Pathogenic *E. coli* is commonly identified as a cause of both acute and persistent paediatric diarrhoea in low- and middle-income countries (31-34), particularly in malnourished children (35, 36). Further, GEMS found that diarrhoea from typical EPEC and heat stable ETEC infections were associated with malnutrition and an increased risk of death in infants (34). There is evidence of chronic carriage or reinfection with EAEC in the birth cohort. While persistent EAEC infections are known to be implicated in long-term diarrhoeal disease in children, this was not evident in our study (37-39). Children diagnosed with DWSD or dysentery in hospital-based surveillance had a high prevalence of DEC, *Campylobacter* spp. and/or *Shigella*, pathogens previously implicated with these conditions (40).

Despite high prevalence of certain pathogens, we did not detect any significant relationships between individual pathogens and wasting or diarrhoeal disease presentation. This may indicate that these outcomes are due to multiple pathogen interactions or additional nutritional or environmental factors. Pathogen virulence likely also plays an important part in the distribution of pathogens between malnourished and/or diarrhoeal children versus those that are relatively healthy (41).

Most pathogens were recovered from faecal samples with limited or no evidence of diarrhoeal disease. While diarrhoeal disease is implicated in child malnutrition, subclinical infections with enteric pathogens may be more strongly linked to poor growth outcomes (12). The MAL-ED study detected high prevalence of *Campylobacter* from an early age and a negative association between high pathogen load and growth, particularly in asymptomatic cases (12). Carriage of enteric pathogens like EAEC and *Campylobacter* can lead to enteric and systemic inflammation and malabsorption of nutrients resulting in malnutrition and faltered growth, particularly in prolonged cases or repeat infections (42-44). Detection of a wide range of pathogens in the absence of diarrhoeal symptoms warrants further investigation to understand the relationship between enteric pathogens and malnutrition (29).

Within Timor-Leste and neighbouring Southeast Asian countries, there is limited evidence on the prevalence of enteric pathogens both in clinical and environmental settings. A multisite study of children aged under five years conducted in Indonesia between 2009 and 2012 detected a high prevalence of rotavirus (37.4%), adenovirus (34.9%) and ETEC (10.0%) (45). Another Indonesian case-control study of diarrhoeal disease conducted between 1997 and 1999 detected very low levels of enteric pathogens in paediatric patients using culture-based methods (i.e., less than 8.0% prevalence for total pathogens)(46), however, it is worth noting that culture-based methods can be lower in sensitivity than PCR-based methods. Our present study aimed to explore environmental and zoonotic pathogen sources in addition to risk factors in the home. Flooding is common in the region, particularly Timor-Leste. This combined with ubiquitous pit latrines and improper garbage disposal provides ample opportunities for transmission of enteric pathogens in the environment. This is supported by a Nigerian study that tested groundwater sources for enteric pathogens. High levels of bacteria including *E. coli, Salmonella, Shigella*, and *Campylobacter* spp. were found in boreholes that likely weren’t adequately maintained and had contamination from nearby septic tanks, sludge systems, and waste dumpsites (47). An Egyptian study also isolated these bacteria from ground water in rural areas (48). While efforts to improve personal practice with water, sanitation, and hygiene measures, food sources and preparation, and animal husbandry are important, exploring the endemicity of enteric pathogens in the environment should also be explored.

There are several limitations of our study. This study was conducted to examine acceptability for undertaking a larger birth cohort study. It was under-powered to examine some important relationships. Timor-Leste is prone to flash-flooding making it difficult to undertake home visits and, on a few occasions, HNGV and NHL experienced flash-flooding. We had reasonably high attrition of our birth cohort, due to highly mobile families especially during the COVID-19 pandemic when the study was interrupted between April and July 2020, leading to substantial loss to follow up. Limited animal faecal samples were able to be collected due an outbreak in African Swine fever in the first six months of the study. In addition, multiplex PCR testing is very sensitive and detects presence of pathogen nucleic acid but not necessarily viable pathogen. The Biofire GI PCR panel has shown high sensitivity and specificity in detection of diarrhoeal pathogens (49). We intended to culture all human clinical *Salmonella, Campylobacter* and *Shigella* samples at NHL, however due to a lack of available resources this was not completed in most cases. In the absence of culture results to compare to the positive PCR results for *Campylobacter, Shigella* and *Salmonella* we were unable to determine the validity of the PCR results in this setting. However, similar studies have shown that multiplex PCR testing can provide an accurate description of enteric pathogen prevalence in low- and middle-income countries (50, 51).

In this study, we demonstrated the possibility of undertaking longitudinal field research in Timor-Leste and produced useful and practical information that will be used to assist in guiding the strategy and approach of future research. Nutrition awareness, choices and availability, food and water safety and exposure to animal and environmental pathogens affect lives of Timor-Leste families and contribute to the multiple malnutrition risk factors. We recommend conducting a larger-scale study to investigate infant and child dietary practices, food safety, WASH, gender, cultural and environmental hygiene conditions in relation to One Health and food safety and security, with a focus on the risk of enteric pathogens. This project should comprehensively investigate malnutrition and stunting risk factors, including zoonotic transmission of enteric pathogens to provide an evidence base for targeted interventions in urban and rural settings in Timor-Leste, utilising a One Health collaborative approach to enhance collaboration and capacity.

## Data Availability

All relevant data are within the manuscript and its Supporting Information files.

## Acknowledgements

The authors thank all persons and organisations that provided support for this study. These include: Professor Nélson Martins and the National Institute of Health-Research Ethics & Technical Committee; Dr Dongbao Yu and Dr Rajesh Pandav at the World Health Organization; Dr Merita Monteiro at Departamentu Kontrolu Moras Hada’et (CDC); Maria Angela Varela Niha at Departamentu Vijilansia Epidemiolojia, Ministério da Saúde; Dr Jeremy Beckett at Maluk Timor; local research team members Francisca Soares and Antoninho Gusmão; Menzies staff including Dr Jennifer Yan, Salvador Amaral, Dr Ian Marr, Lucsendar Alves, Dr Jo Wapling, Karen Champlin, and Dr Shawn Ting; Ismael Barreto and all staff from the National Health Laboratory; all staff from the Diagnostics Laboratory for Animal Health; all staff at HNGV including Dr Flavio Brandão de Araujo, Dr Ingrid Bucens, Dr Amelia Ferreria Pinto and the HNGV Paediatrics team; Ms Melanie McVean and the St John of God Midwifery team; the HNGV Obstetrics and Midwifery team, and; Dr Anna Okello, Dr Francette Geraghty-Dusan, and Bethany Lees from the Australian Centre for International Agricultural Research.

## Supporting Information

**S1 Table. Weight-for-height, height-for-age, and weight-for-age z-score for birth cohort infants at each home visit in Dili, Timor-Leste, 2019-2020**.

**S2 Table. Adjusted univariate odds ratios using a generalised estimating equations model for risk factors associated with pathogen detection for infants in a birth cohort, Dili, Timor-Leste, 2019-2020**.

**S3 Table. Adjusted univariate odds ratios using a generalised estimating equations model for risk factors associated with moderate wasting for infants in a birth cohort, Dili, Timor-Leste, 2019-2020**.

**S4 Table. Mean age in months at admission by diagnosis and type of enteric pathogen detected for hospital-based surveillance cases, Dili, Timor-Leste, 2019–2020**.

**S5 Table. Adjusted univariate odds ratios using a generalised estimating equations model for pathogens and host factors associated with hospitalisation for infants in a birth cohort, Dili, Timor-Leste, 2019-2020**.

